# Novel Application of a Force Sensor during Sit-to-Stands to Measure Dynamic Cerebral Autoregulation Onset

**DOI:** 10.1101/2022.01.14.22269108

**Authors:** Alicen A. Whitaker, Eric D. Vidoni, Stacey E. Aaron, Adam G. Rouse, Sandra A. Billinger

**Author notes:** **Corresponding Author:** Sandra A. Billinger, PT, PhD; Address: 3901 Rainbow Blvd, Mail Stop 2002, Kansas City, KS 66160. **Author Contributions:** AW, EV, SB conceived and designed research; AW and SA performed experiments; AW, EV, SA, SB analyzed data; AW, EV, SA, AR, SB interpreted results of experiments; AW, EV, SA, AR, SB drafted manuscript; AW, EV, SA, AR, SB edited and revised manuscript; AW, EV, SA, AR, SB approved final version of manuscript; AW prepared figures.

## Abstract

**Purpose:** Current sit-to-stand methods measuring dynamic cerebral autoregulation (dCA) do not capture the precise onset of the time delay (TD) response. Reduced sit-to-stand reactions in older adults and individuals post-stroke could inadvertently introduce variability, error, and imprecise timing. We applied a force sensor during a sit-to-stand task to more accurately determine how TD before dCA onset may be altered.

**Methods:** Middle cerebral artery blood velocity (MCAv) and mean arterial pressure (MAP) were measured during two sit-to-stands separated by 15 minutes. Recordings started with participants sitting on a force-sensitive resistor for 60 seconds, then asked to stand for two minutes. Upon standing, the force sensor voltage immediately dropped and marked the exact moment of arise-and-off (AO). Time from AO until an increase in cerebrovascular conductance (CVC = MCAv/MAP) was calculated as TD.

**Results:** We tested the sensor in 4 healthy young adults, 2 older adults, and 2 individuals post-stroke. Healthy young adults stood quickly and the force sensor detected a small change in TD compared to classically estimated AO, from verbal command to stand. When compared to the estimated AO, older adults had a delayed measured AO and TD decreased up to ~53% while individuals post-stroke had an early AO and TD increased up to ~14%.

**Conclusion:** The transition reaction speed during the sit to stand has the potential to influence dCA metrics. As observed in the older adults and participants with stroke, this response may drastically vary and influence TD.

**New & Noteworthy:** We developed a force sensor and are the first to apply it during sit-to-stands measuring the dynamic cerebral autoregulation (dCA) response. Current methodologies estimate the transition reaction speed of the sit-to-stand, which influences the dCA metrics. Compared to estimating the kinetic moment of arise-and-off (AO), our force sensor can detect early or delayed AO in individuals who may present with lower extremity weakness such as older adults and clinical populations such as stroke.

## Introduction

The brain requires constant cerebral blood flow, even during perturbations to the systemic circulation.(1) Dynamic cerebral autoregulation (dCA) is the ability of the brain to maintain blood flow by altering cerebrovascular conductance (CVC) during perturbations to circulation, such as changes in peripheral blood pressure.(2-4) Recording the dCA response has become prevalent with the use of transcranial Doppler ultrasound to measure middle cerebral artery blood velocity (MCAv) and finger plethysmography to measure mean arterial pressure (MAP).(4, 5) During a transient drop in peripheral blood pressure, for example in the transition from a seated position to standing, the time delay (TD) before the onset of the dCA response is measured as the elapsed time before an increase in CVC.(6, 7)

A sit-to-stand transition is a common experimental perturbation utilized to measure the temporal dCA response.(8) When performing a sit-to-stand, peripheral blood pressure transiently drops due to gravitational venous pooling and vasodilation.(9) To counteract the drop in blood pressure, dCA responds by increasing the conductance of MCAv to the cerebrovascular system before MAP increases in the peripheral vascular system.(6)

While previous studies in healthy young and older adults have performed the sit-to-stand technique to measure the dCA response,(6, 8, 10-15) the exact methods of characterizing dCA have varied between studies. While some report having the individual stand quickly (within 0-3 seconds) and mark the initiation of standing as when the verbal command was given,(6, 10-12) others report having the individual place their feet on a stool in front of them (legs at 90 degrees) and mark the initiation of standing from when their feet touch the floor.(8, 13-15) However, the moment of standing from a seated position is not objectively marked.(16) The start of the postural change, the stimulus which drops peripheral blood pressure, is critical for measuring the timing of the dCA response and has been indeterminate in prior protocols. Considering the sit-to-stand transition influences hemodynamics, not measuring the exact moment could inadvertently introduce variability, error, and imprecise timing of the dCA metrics. Measuring the exact moment of stance is especially important in older adults and clinical populations, such as stroke who may present with reduced lower extremity strength, resulting in a delayed sit-to-stand reaction.(17-19) Current methodologies are unable to account for the potential delay between the initiation and the stance phase of a sit-to-stand.(16)

To address this gap in the literature, we developed a force sensor to determine the kinetic moment of arise-and-off (AO).(16) Compared to estimating AO from a verbal command similar to previous work, (6, 10-12) we hypothesized that measuring the exact moment of AO, measured with a force sensor, would alter the calculation of the TD before the onset of the dCA response. We conducted the procedure twice separated by 15 minutes (T1= first sit to stand and T2 = second sit to stand).

## Methods

This is an ongoing study, “Blood Flow Response and Acute Interval Exercise (BRAIN), focused on the characterization of the cerebrovascular hemodynamic response to a single bout of high intensity interval exercise and the sit-to-stand dCA response before and after exercise (NCT04673994). We developed the force sensor prior to study initiation and present limited data to establish “proof of principle,” not as interim analyses of the a priori stated primary outcome of the clinical trial. We recruited and enrolled healthy young adults, older adults, and individuals post-stroke. Healthy young adults were included if they were between the ages of 18-30 years old and classified as low cardiovascular risk using the American College of Sports Medicine (ACSM) Cardiovascular Risk Screen.(20) Older adults and individuals post-stroke (6 months to 5 years post-stroke) were included if they were: 1) between the ages of 40-80 years old, 2) performed less than 150 minutes of moderate intensity exercise per week, and 3) able to answer consenting questions and follow a 2-step command. Individuals were excluded if they 1) were unable to stand from a sitting position without physical assistance, 2) had a previous history of another neurological disease (i.e. multiple sclerosis, Alzheimer’s disease, or Parkinson’s disease), and 3) did not have a MCAv signal on the right or left temporal window using transcranial Doppler ultrasound (TCD). The study was approved by the University of Kansas Medical Center Institutional Review Board Human Subjects Committee.

Prior to study procedures, all participants provided written informed consent to participate and we collected demographic information. Participants also completed a standardized 5 time sit-to-stand assessment, to determine leg strength and balance by measuring how quickly an individual could stand up from a chair and sit down 5 times without the use of their arms.(17, 21, 22)

### Set Up

Consistent with previously published protocols, our laboratory room was dimly lit and kept at a controlled temperature of 22°C to 24°C.(23) Prior to the visit, participants were asked to abstain from caffeine for 8 hours,(24-26) vigorous exercise for 24 hours,(27) and alcohol for 24 hours.(28) Participants were seated on a recumbent stepper (T5XR NuStep, Inc. Ann Arbor, MI) with the chair rotated 90 degrees perpendicular. The participant sat with feet flat on the ground, legs at 90 degrees, and an upright trunk posture. The participants rested for 20 minutes while the equipment was donned: 1) a 5-lead electrocardiogram (ECG; Cardiocard, Nasiff Associates, Central Square, New York) to measure heart rate, 2) a left middle finger plethysmograph (Finometer, Finapres Medical Systems, Amsterdam, the Netherlands) to measure beat-to-beat MAP, and 3) bilateral TCD probes (2-MHz, Multigon Industries Inc, Yonkers, New York) to measure MCAv. The TCD probes were secured with headgear to maintain the optimal position and angle. Consistent with previously published methodology measuring beat-to-beat MAP during a sit-to-stand, participants were instructed to place their left hand with the finger plethysmograph flat on the upper chest at heart level.(6, 8, 13) Due to neuromuscular fatigue, an arm sling was used to help hold the Finometer in place during the sit-to-stand recording.(13) For individuals with upper extremity spasticity, the Finometer was placed on the non-affected upper extremity.

### Force-Sensor

Our custom force sensor was comprised of a force-sensitive resistor (SEN-09376, Mouser Electronics Inc., Tx) connected to an operational amplifier (Canaduino LM393, Universal-Solder, Ontario, Canada) and custom circuit housed on a breadboard (Inland 400 tie-point), shown in **Figure 1**. The force-sensitive resistor was placed on the seat under the visualized right ischial tuberosity and connected to the custom circuit by 6 feet of stranded wire. For individuals with stroke, the force-sensitive resistor was placed underneath the non-affected lower extremity due to asymmetrical weight bearing during a sit-to-stand.(29) When sitting, the force-sensitive resistor converted the weight of the participant into a voltage, amplified at the breadboard, and digitized via an analog-to-digital unit (NI-USB-6212, National Instruments). Once participants stood from the chair the force-sensitive resistor no longer detected the participant’s weight and the voltage signal quickly decreased to 0 volts for the remainder of the recording. The voltage of our force sensor was captured simultaneously with MCAv and MAP during the sit-to-stand using custom written software. All measures were recorded at 500 Hz using a custom written software within MATLab implementing the Data Acquisition Toolbox (R2019a, TheMathworks Inc, Natick, Massachusetts).

**Figure 1.**
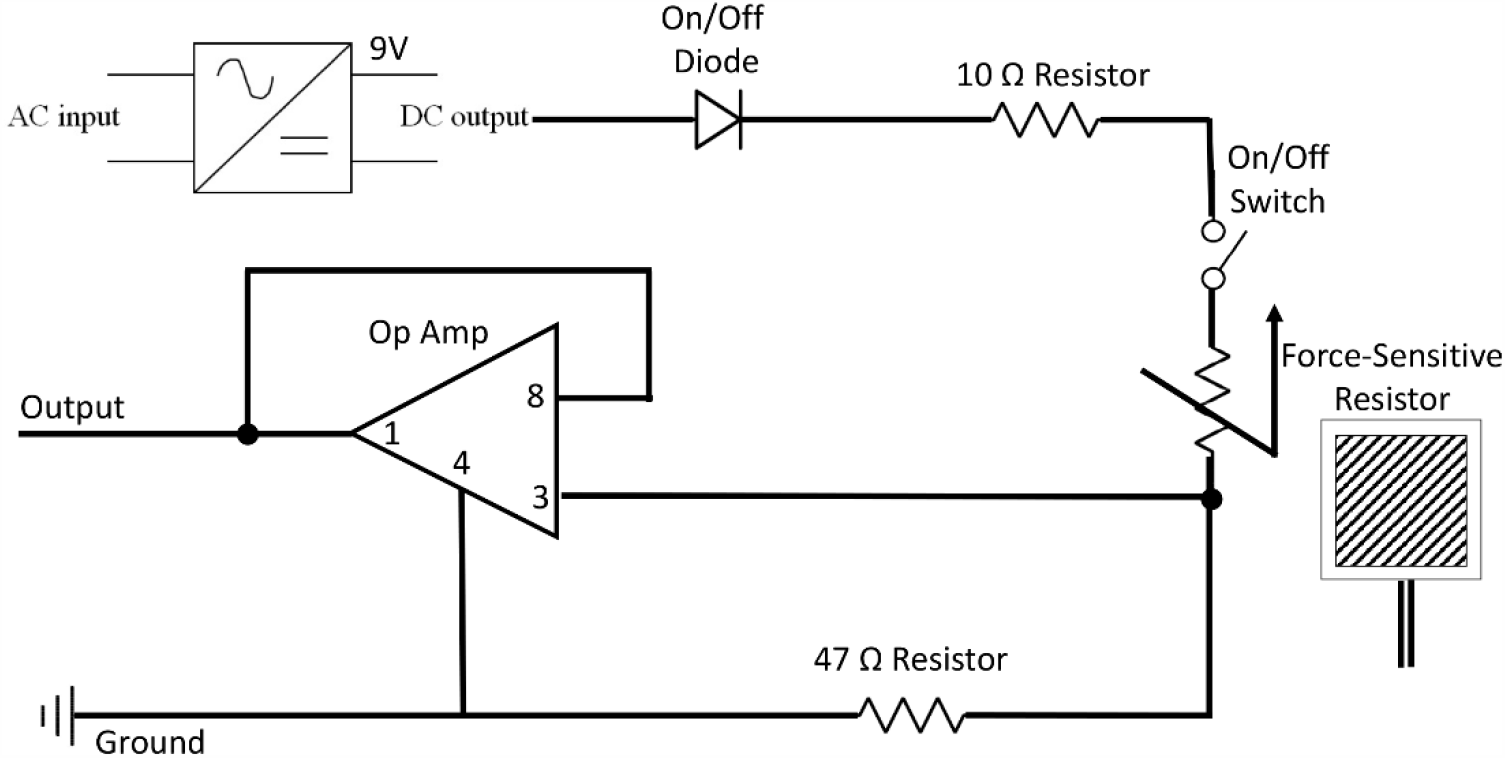
Drawing of the Custom Force Sensor Measuring AO during Sit-to-Stands. For the operational amplifier (Op Amp), the pins correspond to: 8-Vcc, 3-Input, 1-Output, 4-Ground. The force-sensitive resistor has a resistance that decreases with force. As the subject stands, force decreases, the resistance increases, and the voltage drop increases at the force-sensitive resistor. This results in a decreased Op Amp input voltage due to decreased voltage drop across the 47 Ohm resistor.

### Sit-to-Stand

Participants were familiarized with the sit-to-stand protocol prior to the recording. After a minute of seated rest, participants were given a three-second countdown and asked to stand at 60 seconds into the recording. Participants remained standing for two minutes to allow hemodynamic stabilization.(6, 10) The sit-to-stand recording was performed twice separated by 15 minutes (T1, T2).

Raw data were processed offline using the QRS complex within the ECG to calculate beat-to-beat MCAv and MAP.(12) AO was defined as the minimum of the second derivative of the sensor voltage, corresponding with the acceleration of the decreasing voltage velocity during the sit-to-stand.

### Data Analysis

Based on previously published methodology, the TD before the onset of the dCA regulatory response was measured as seconds between performing the sit-to-stand and a continuous increase in CVC without transient reduction (CVC = MCAv/MAP).(6, 10) Rather than estimating AO in the recording as 60 seconds, our force sensor marks the exact AO time. We calculated the difference in the TD response when using the force sensor compared to estimating AO at 60 seconds, or when the participant was asked to stand.

## Results

A total of 8 individuals completed the sit-to-stand transition with the force sensor to measure the TD before the onset of dCA. We included healthy young adults (n=4), older adults (n=2) and individuals post-stroke (n=2). Participant characteristics are shown in **Table 1**. The sit-to-stand AO time for all participants is shown in **Table 2**.

**Table 1.** Participant Characteristics.

| Subject | Age Range (years) | Sex | 5 Time Sit to Stand (seconds) |
| --- | --- | --- | --- |
| Young Adults |  |  |  |
| 1 | 21 - 25 | F | 5.05 |
| 2 | 21 - 25 | F | 7.42 |
| 3 | 21 - 25 | M | 6.42 |
| 4 | 26 - 30 | M | 5.39 |
| Older Adults |  |  |  |
| 5 | 66 - 70 | M | 6.54 |
| 6 | 71 - 75 | M | 10.85 |
| Individuals Post-Stroke |  |  |  |
| 7 | 41 - 45 | M | 10.07 |
| 8 | 71 - 75 | M | 26.35 |

**Table 2.**
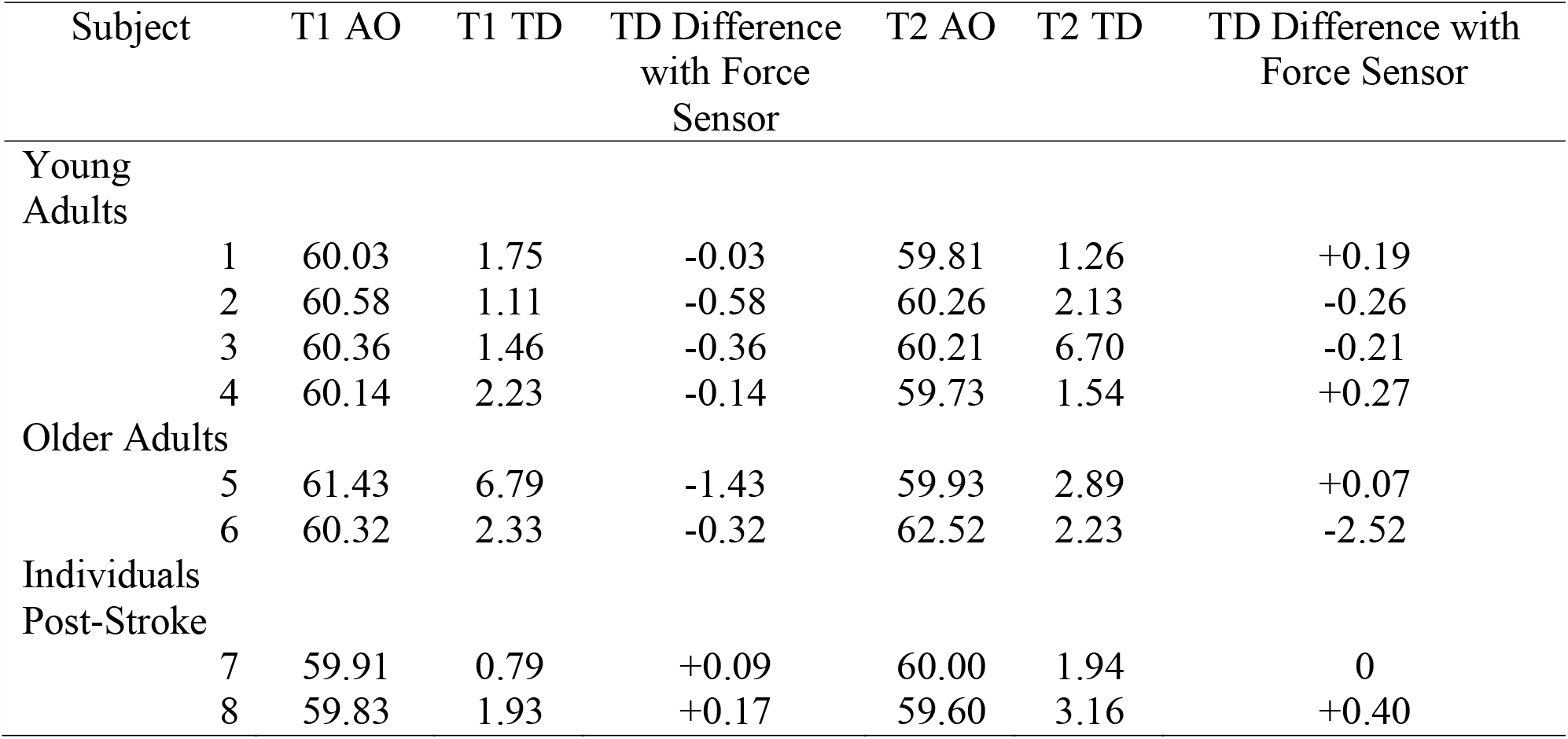
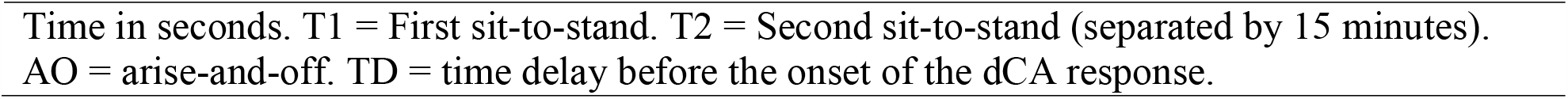
Sit-to-Stand Outcomes.

| Subject | T1 AO | T1 TD | TD Difference with Force Sensor | T2 AO | T2 TD | TD Difference with Force Sensor |
| --- | --- | --- | --- | --- | --- | --- |
| Young Adults |  |  |  |  |  |  |
| 1 | 60.03 | 1.75 | -0.03 | 59.81 | 1.26 | +0.19 |
| 2 | 60.58 | 1.11 | -0.58 | 60.26 | 2.13 | -0.26 |
| 3 | 60.36 | 1.46 | -0.36 | 60.21 | 6.70 | -0.21 |
| 4 | 60.14 | 2.23 | -0.14 | 59.73 | 1.54 | +0.27 |
| Older Adults |  |  |  |  |  |  |
| 5 | 61.43 | 6.79 | -1.43 | 59.93 | 2.89 | +0.07 |
| 6 | 60.32 | 2.33 | -0.32 | 62.52 | 2.23 | -2.52 |
| Individuals Post-Stroke |  |  |  |  |  |  |
| 7 | 59.91 | 0.79 | +0.09 | 60.00 | 1.94 | 0 |
| 8 | 59.83 | 1.93 | +0.17 | 59.60 | 3.16 | +0.40 |

Time in seconds. T1 = First sit-to-stand. T2 = Second sit-to-stand (separated by 15 minutes). AO = arise-and-off. TD = time delay before the onset of the dCA response.

### Healthy Young Adults

At T1, the AO time for the healthy young adults was 60.28 ± 0.24 seconds. The force sensor was able to account for a shorter TD by 0.28 ± 0.24 seconds compared to estimating AO from the verbal command to stand. At T2, the AO time was on average at 60.00 ± 0.27 seconds in the recording. However, because 2 individuals had an AO time before being asked to stand, the force sensor accounted for a change in the TD response of 0.003 ± 0.27 seconds.

### Older Adults

At T1, participant #5’s AO time was 61.43 seconds. Compared to estimating AO from the verbal command to stand, the force sensor was able to account for a decrease in the TD response by 1.43 seconds (~17%). At T2, the same participant #5 had a 1.50 second faster AO time compared to T1 and had an AO time before being asked to stand. In contrast, while participant #6 had a small change in the TD at T1 of 0.32 seconds, they were 2.20 seconds slower at T2. The force sensor was therefore able to account for a decrease in the TD response by 2.52 seconds (~53%) at T2, compared to estimating AO from the verbal command to stand (shown in **Figure 2**).

**Figure 2.**
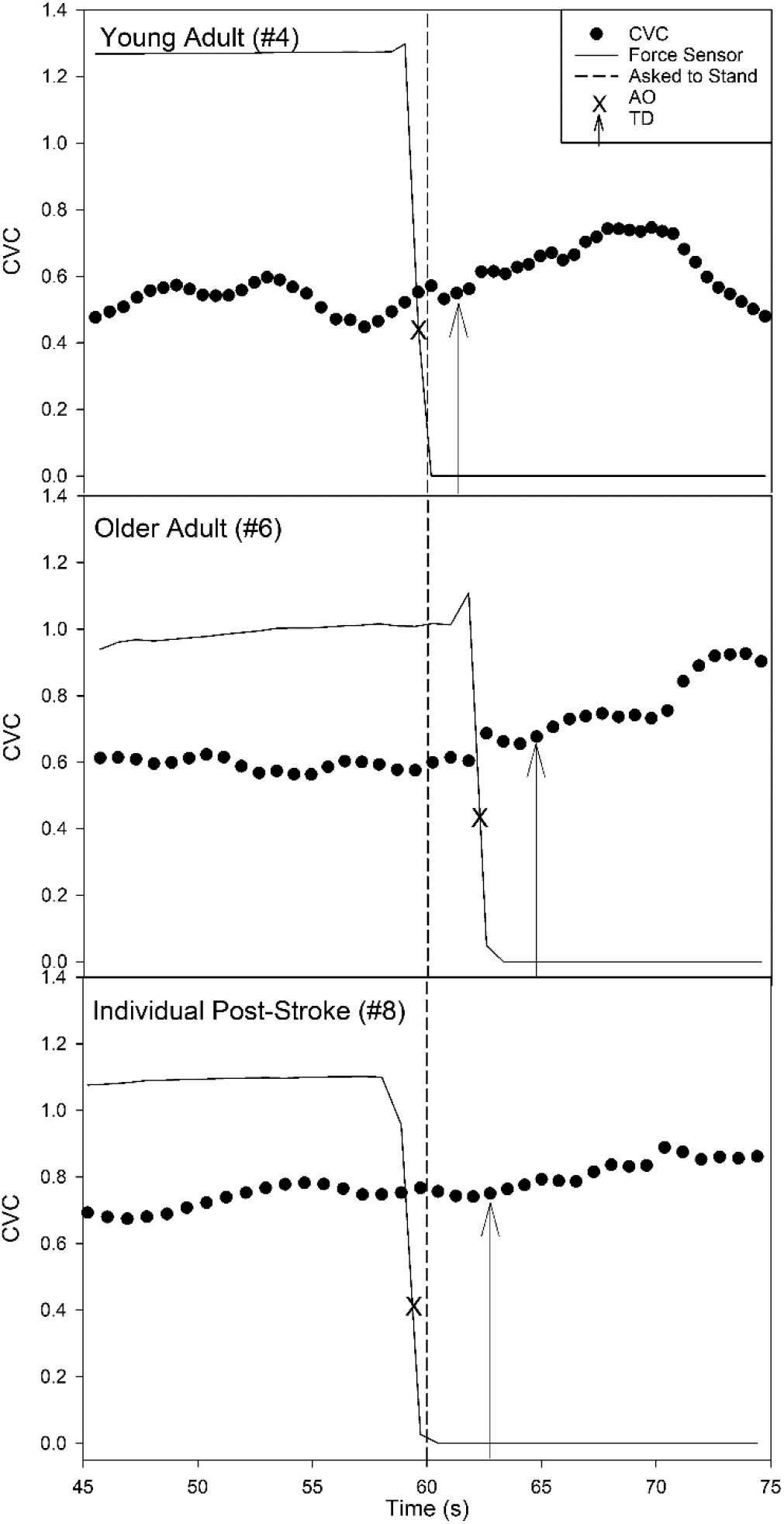
Novel Application of a Force Sensor during a Sit-to-Stand in a Single Healthy Young Adult (n = 1), Older Adult (n = 1), and Individual Post-Stroke (n = 1). CVC = Cerebrovascular Conductance. Horizontal Dotted Line = Individuals were asked to perform the sit-to-stand at 60 seconds. AO = Arise-and-off when participants stood from the chair. TD = Time delay before the onset of dynamic cerebral autoregulation. Force sensor values are in arbitrary units.

### Individuals Post-Stroke

At T1, participant #7’s AO time was 0.09 seconds before the verbal command to stand, which accounted for an increase (~13%) in the TD response, compared to estimating AO from the verbal command to stand. At T2, the same participant #7’s AO time was at exactly 60 seconds. In contrast, participant #8’s AO was before the verbal command to stand, which accounted for an increase in the TD response at T1 (~10%) and T2 (~14%), respectively.

## Discussion

Previously published sit-to-stand methodology measuring the TD before the onset of the dCA response does not report the true moment of AO from the chair, and therefore does not measure the exact start of the stimulus that drops peripheral blood pressure. While force sensors have been previously used in the kinetic characterization of a sit-to-stand,(30) we are the first to apply a force sensor during a hemodynamic recording of dCA onset. The force sensor allowed for the simultaneous measurement of the sit-to-stand reaction behavior with the physiologic response.

We hypothesized that measuring AO precisely with a force sensor would identify previously unobserved variability in standing dynamics compared to estimating AO from a verbal command or other methods.(6, 8, 10, 13) We have shown that while healthy young adults may demonstrate a faster transition reaction speed during a sit-to-stand dCA recording,(4, 8) older adults and individuals post-stroke may differ in their AO response. We have shown that the TD before the onset of dCA, which typically occurs within 10 seconds,(6, 8, 10, 14) could potentially be skewed in some individuals by more than 2.5 seconds, which would directly affect dCA metrics.

Previous reviews have also reported delayed sit-to-stand reaction time in individuals with stroke,(18, 31, 32) which was supported by our results (see Table 1) reporting the 5 time sit-to-stand scoring 2 standard deviations away from the age-normative values.(19) While this would suggest an individual post-stroke would have a delay in AO, our results show that the individuals with stroke began to stand prematurely during the 3 second countdown, shown in **Figure 2**. A possible explanation for an early AO could be that the individuals post-stroke anticipated longer sit-to-stand transition times.(17)

We show that the healthy young adult participants performed AO at approximately the same time between T1 and T2. In fact, 2 healthy young adults stood early at T2, which could have been due to anticipation and learning affect. One older adult participant could have also had an increase in performance learning and stood faster at T2. However, the other older adult took ~2 seconds longer to perform AO at T2, which may have been due to lower extremity weakness. The individuals post-stroke may have recognized their lower extremity weakness and anticipated needing longer to stand.

Imprecise temporal measures of dCA could potentially impact our interpretation of data especially when comparing healthy individuals to clinical populations. A previous study performed repeated sit-to-stands every 10 seconds in healthy older adults compared to individuals with dementia and mild cognitive impairment.(33) Contrary to their hypothesis, they reported individuals with mild cognitive impairment and dementia had a better autoregulation response.(33) However, the individuals with cognitive impairment could have performed the sit-to-stands early and, therefore, could have skewed the response. Without measuring the exact start of the sit-to-stand stimulus, the wrong conclusions could unintentionally be drawn on the temporal dCA response. Another example is a study comparing the sit-to-stand dCA response after a 14-week exercise intervention in pregnant women that reported no significant change in the TD before the onset of dCA.(34) However, there could have been a large difference in the sit-to-stand response between the baseline measures taken during the 2^nd^ trimester of pregnancy and measures taken after the exercise intervention during the 3^rd^ trimester of pregnancy. Pregnant women within the 3^rd^ trimester could have had a slower sit-to-stand time that was not accounted for and could have inadvertently skewed the calculation of TD. Imprecise timing of the dCA response, supported by our results showing ~10% to 53% change in TD, could systematically impact study findings.

## Conclusions

The implementation of a force sensor to measure AO during a hemodynamic sit-to-stand recording could improve the exact calculation of the TD before the onset of the dCA response. Applying the force sensor into the sit-to-stand dCA methodology has the potential to improve scientific rigor and reproducibility, especially in older adults and clinical populations such as stroke.

## Supporting information

coi_aaron

coi_billinger

coi_rouse

coi_vidoni

coi_whitaker

## Data Availability

All data produced in the present study are available upon reasonable request to the authors.

## Acknowledgements

We would like to thank Preston Judd for his contributions to editing and proof reading.

